# Risk factors for SARS-CoV-2 infection, hospitalisation, and death in Catalonia, Spain: a population-based cross-sectional study

**DOI:** 10.1101/2020.08.26.20182303

**Authors:** Judit Villar-García, Rosa María Vivanco-Hidalgo, Montse Clèries, Elisenda Martinez, David Monterde, Pol Perez-Sust, Luis Garcia-Eroles, Carol Sais, Montse Moharra, Emili Vela

**Author notes:** Correspondence to: Judit Villar-García. Equally contribution (Judit Villar-García, Rosa María Vivanco-Hidalgo).

## Abstract

**OBJECTIVE:** To identify the different subpopulations that are susceptible for severe acute respiratory syndrome coronavirus 2 (SARS-CoV-2) infection and hospitalisation or death due to coronavirus disease 2019 (COVID-19) in Catalonia, Spain.

**DESIGN:** Cross-sectional study.

**SETTING:** Data collected from the Catalan Health Surveillance System (CatSalut) in Catalonia, a region of Spain.

**PARTICIPANTS:** Using data collected between 1 March and 1 June 2020, we conducted the following comparative analyses: people infected by SARS-CoV-2 (328 892) vs Catalonia’s entire population (7 699 568); COVID-19 cases who required hospitalisation (37 638) vs cases who did not require hospitalisation (291 254); and COVID-19 cases who died during the study period vs cases who did not die during the study period (12 287).

**MAIN OUTCOME MEASURES:** Three clinical outcomes related to COVID-19 (infection, hospitalisation, or death). We analysed sociodemographic and environment variables (such as residing in a nursing home) and the presence of previous comorbidities.

**RESULTS:** A total of 328 892 cases were considered to be infected with SARS-CoV-2 (4.27% of total population). The main risk factors for the diagnostic were: female gender (risk ratio [RR] =1.49; 95% confidence interval [95% CI] =1.48-1.50), age (45-64 years old; RR=1.02; 95% CI=1.01-1.03), high comorbidity burden (GMA index) (RR=3.03; 95% CI=2.97-3.09), reside in a nursing home (RR=11.82; 95% CI=11.66-11.99), and smoking (RR=1.06; 95% CI=1.05-1.07). During the study period, there were 37 638 (11.4 %) hospitalisations due to COVID-19, and the risk factors were: male gender (RR=1.45; 95% CI=1.43-1.48), age > 65 (RR=2.38; 95% CI=2.28-2.48), very low individual income (RR=1.03; 95% CI=0.97-1.08), and high burden of comorbidities (GMA index) (RR=5.15; 95% CI=4.89-5.42). The individual comorbidities with higher burden were obesity (RR=1.23; 95% CI=1.20-1.25), chronic obstructive pulmonary disease (RR=1.19; 95% CI=1.15-1.22), heart failure (RR=1.19; 95% CI=1.16-1.22), diabetes mellitus (RR=1.07; 95% CI=1.04-1.10), and neuropsychiatric comorbidities (RR=1.06; 95% CI=1.03-1.10). A total of 12 287 deaths (3.73%) were attributed to COVID-19, and the main risk factors were: male gender (RR=1.73; 95% CI=1.67-1.81), age > 65 (RR=37.45; 95% CI=29.23-47.93), residing in a nursing home (RR=9.22; 95% CI=8.81-9.65), and high burden of comorbidities (GMA index) (RR=5.25; 95% CI=4.60-6.00). The individual comorbidities with higher burden were: heart failure (RR=1.21; 95% CI=1.16-1.22), chronic kidney disease (RR=1.17; 95% CI=1.13-1.22), and diabetes mellitus (RR=1.10; 95% CI=1.06-1.14). These results did not change significantly when we considered only PCR-positive patients.

**CONCLUSIONS:** Female gender, age between 45 to 64 years old, high burden of comorbidities, and factors related to environment (nursing home) play a relevant role in SARS-CoV-2 infection and transmission. In addition, we found risk factors for hospitalisation and death due to COVID-19 that had not been described to date, including comorbidity burden, neuro-psychiatric disorders, and very low individual income. This study supports interventions for transmission control beyond stratify-and-shield strategies focused only on protecting those at risk of death. Future COVID-19 studies should examine the role of gender, the burden of comorbidities, and socioeconomic status in disease transmission, and should determine its relationship to workplaces, especially healthcare centres and nursing homes.

## Introduction

In December 2019, an outbreak of severe acute respiratory syndrome coronavirus 2 (SARS-CoV-2), which causes the so-called coronavirus disease 2019 (COVID-19), occurred in Wuhan, China.^1^ Within six months, this new virus has spread worldwide, affecting 216 countries, and causing more than half a million deaths.^2^ The rapid transmission of the disease, the presence of asymptomatic carriers, and the swift deterioration of some patients have resulted in bursts of COVID-19 cases that have overwhelmed many hospitals, especially intensive care (ICU) units, in many developed countries.^3,4^ However, it is not yet clear which risk factors should be considered when managing COVID-19 from the healthcare and public health perspective.^5^ Therefore, it is important to distinguish the risk factors for infection by SARS-CoV-2 from those for hospitalisation or death due to COVID-19. A UK study using primary care data found that the risk factors for SARS-CoV-2 infection were increasing age, male gender, population density, socioeconomic deprivation, and black ethnicity.^6^ People with a high risk of becoming infected do not always develop a severe disease condition, although they might be a source of infection transmission.^7,8^ To improve the management of this new disease, several studies have sought to identify the risk factors for hospitalisation and death, and found the most common factors to be age, male gender, cardiovascular diseases, chronic obstructive respiratory disease (COPD), obesity, and malignant diseases.^1,9–15^ In contrast, a US study found that older persons, men, and those with preexisting hypertension and/or diabetes had greater risk of hospitalisation.^16^ This risk should be considered carefully because it may cause hospitals to become overloaded during times of epidemic.^3,4^

However, studies attempting to define the characteristics of COVID-19 patients have shown some inconsistencies between countries. Several studies in the Chinese population have reported that COVID-19 patients suffer from cardiovascular diseases, chronic respiratory diseases, and diabetes.^17–19^ In Italy, COVID-19 patients with a concomitant cardiac disease had a poorer prognosis than those without cardiac involvement.^20^ In contrast, several US studies have reported obesity as a recurrent clinical characteristic of COVID-19 inpatients,^16,21,22^ which is likely because obesity is more prevalent in the US (~40%) than in other Western countries (e.g. 20% in Italy and 24% in Spain).^23^ In addition, smoking was a strong risk factor in Chinese COVID-19 patients but was not considered relevant in the US population.^9^ The risk factors for mortality due to COVID-19 are similar to those for hospitalisation, and include increasing age, male gender, and chronic comorbidities, including obesity.^13^ Thus, it is important to determine the risk of death due to COVID-19 in order to identify the groups of patients requiring early implementation of measures for prevention, diagnosis, and treatment.

Therefore, epidemiological studies performed in specific countries could reveal differences that are important for the management of the disease. In this sense, Catalonia was one of the Spanish regions with most cases of COVID-19,^24^ and its particularities may be reflected in the characteristics of its coronavirus patients. Hence, identifying the clinical characteristics of COVID-19 patients in Catalonia may help define risk factor profiles for similar regions. If we can detect early those individuals with the highest risk of becoming infected and transmitting the disease to the vulnerable population, we can decrease the number of hospitalisations and deaths.^24^

In this report, we described the sociodemographic and clinical characteristics of individuals who have been infected or hospitalised, or who died due to COVID-19 in Catalonia, Spain. To attempt to cover the full spectrum of the disease, the aim of our study was to identify the risk factors for becoming infected with SARS-CoV-2 and developing symptoms, the risk factors that are predictive of hospitalisation, and the risk factors that are predictive of death related due to COVID-19.

## Methods

### Study Context and Design

We performed a population-based cross-sectional study of the entire population of Catalonia (7 699 568). We collected data from the Catalan Health Surveillance System (CatSalut), a large, exhaustive, administrative healthcare database that includes information on medical diagnoses, healthcare resource use, and individual income for all Catalan residents. Data from different health-administrative databases were linked and de-identified by a team not involved in the study analysis; the study investigators only had access to a fully anonymized database. The study protocol was approved by the Ethical Review Board of IMIM (Code 2020/9368, Barcelona, Spain). We report this study according to the STROBE guidelines for observational studies using routinely collected data.

The first case of COVID-19 in Catalonia was declared in late February 2020 and lockdown was ordered on March 13, 2020. The peak of incidence was reached in early April 2020, and was followed by a slow progressive decline in the number of incident cases thereafter. We included all consecutive patients declared by the epidemiological surveillance system in the SARS-CoV-2 registry (RSACovid-19), by their primary healthcare facility team, or who were admitted to a hospital (public or private), including emergency admissions, with a diagnosis of SARS-CoV-2 infection between 1 March 2020 and 1 June 2020 inclusive.

### Data collection

Catalonia has a universal public healthcare system, including primary and hospital healthcare, as well as prescription drug costs. The Catalan Central Registry of Insured Persons allows us to link individual-level information between all registries.

We collected information on COVID-19 diagnosis, demographics, admission to nursing homes, and pre-existing comorbidities (hypertension, diabetes mellitus, obesity, hyperlipidaemia, smoking, ischemic heart disease, heart failure, stroke, COPD, HIV-AIDS, chronic kidney disease [CKD], cirrhosis, dementia, malignant neoplasm, mental disability, chronic psychiatric disease (including alcohol dependence disorder), and a comorbidity index [adjusted comorbidity groups, GMA]).^25^ The GMA index is a morbidity measurement recently developed and adapted to the Spanish healthcare system, and that classifies the population into six morbidity groups. The explanatory value of GMA has been verified by comparing it with other morbidity measures such as the Charlson index or the number of chronic diseases. Our data sources included the RSACovid-19 registry (surveillance registry), laboratory registries, and primary care and hospital administrative databases.

### Outcomes

Cases were considered to be infected with SARS-CoV-2 if they were polymerase chain reaction (PCR)-positive, if they had a positive serological test result, or if they had been diagnosed as such by an MD or epidemiologist according to stringent validated diagnostic criteria (current WHO/ECDC criteria).^26,27^ All deaths related to coronavirus were included, whether they were hospitalised or not. The scarcity of PCR tests during the pandemic precluded the testing of all suspected cases of COVID-19. For that reason, we considered infection according to molecular criteria (positive on a PCR or positive a serological test) and/or clinical/epidemiological criteria.

### Statistical analysis

We used frequencies and percentages to describe the categorical variables, and medians and interquartile ranges (IQR) for continuous variables (Table 1). If for a given patient the database/registry did not include information on any comorbidities, we assumed that none were present. We compared baseline demographic and comorbidity data between: individuals considered to be infected with SARS-CoV-2 vs the rest of the population; COVID-19 cases who required hospitalisation vs cases who did not require hospitalisation; and COVID-19 cases who died during the study period vs cases who did not die during the study period.

**Table 1.**
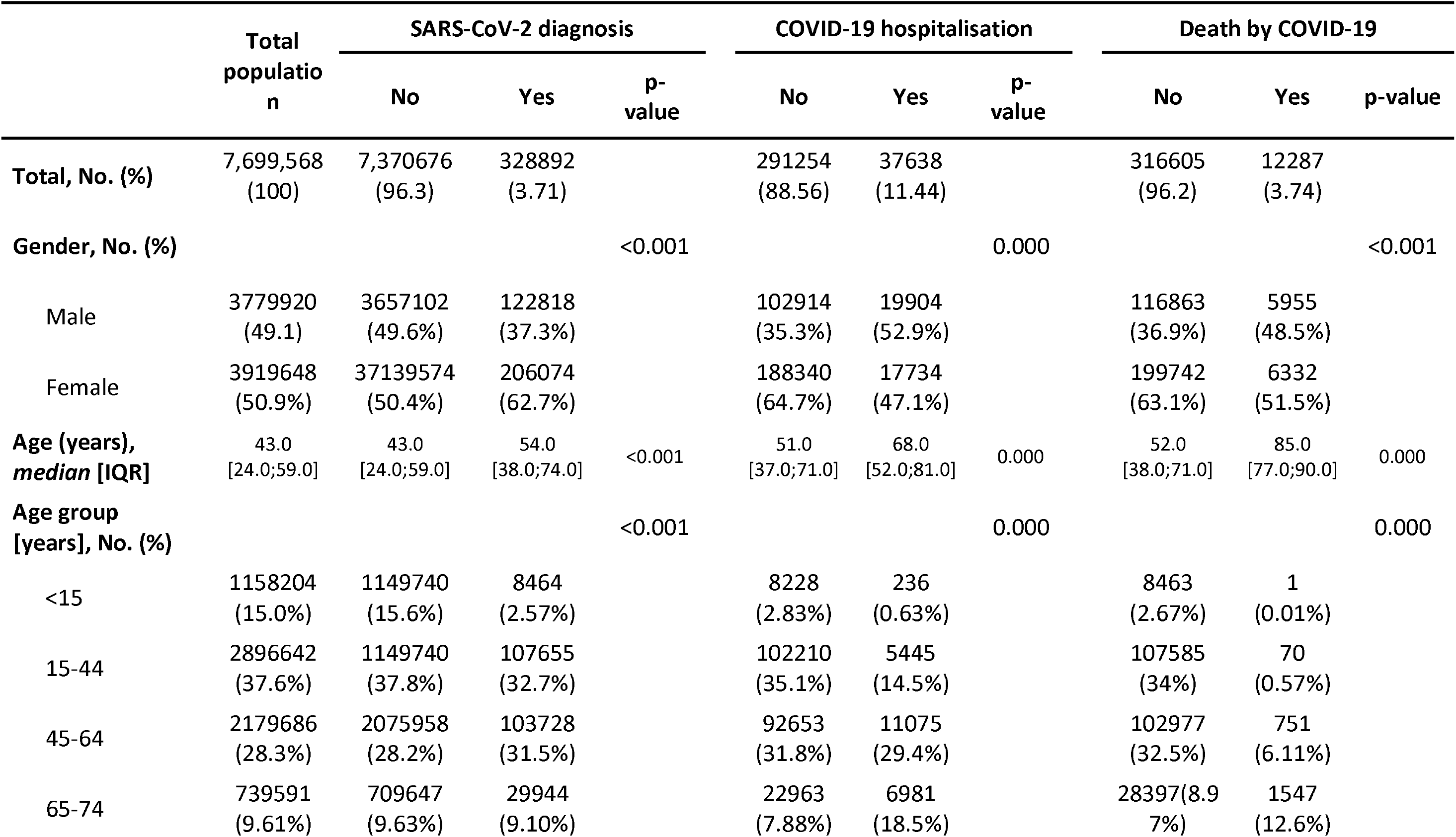
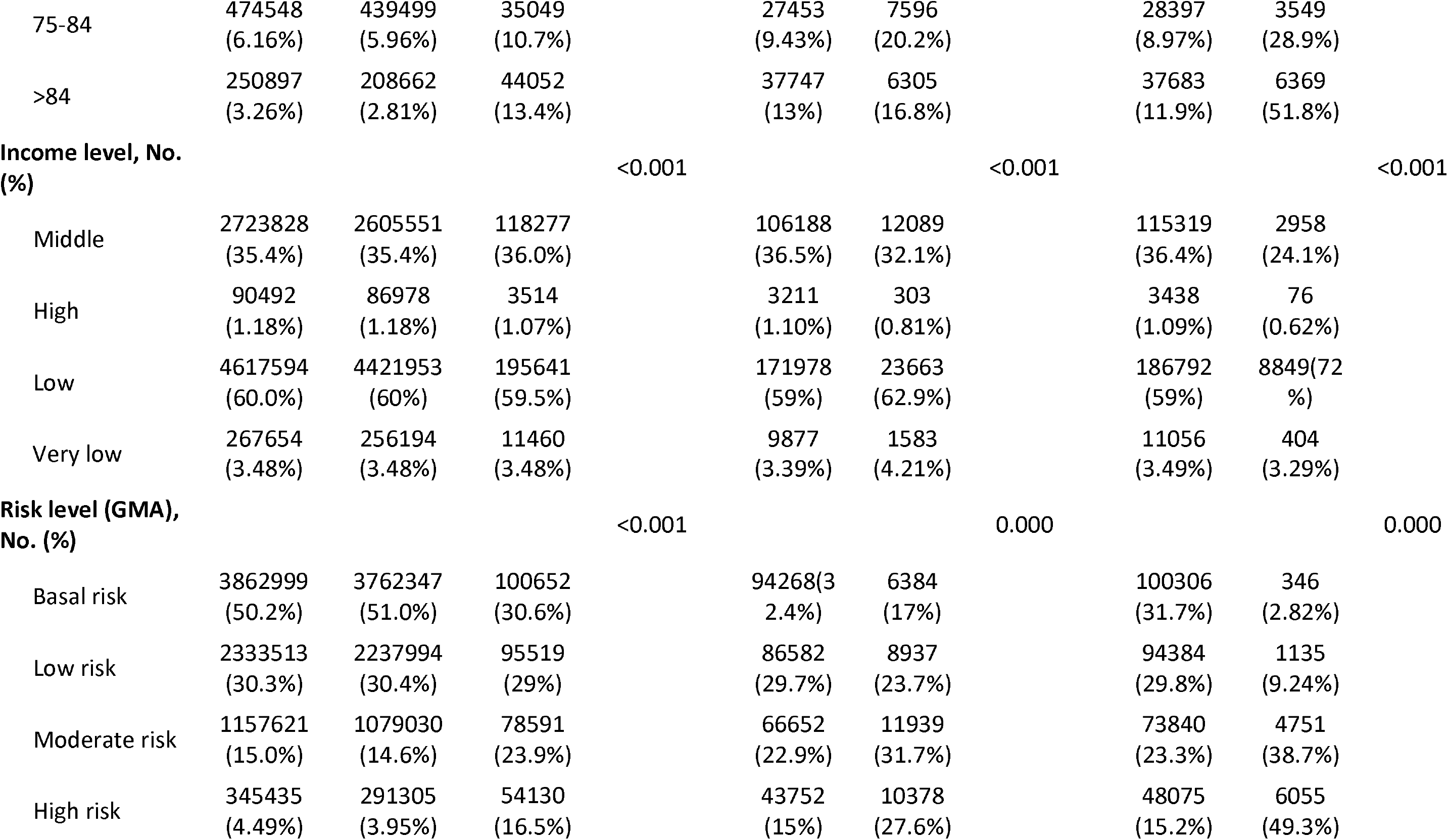
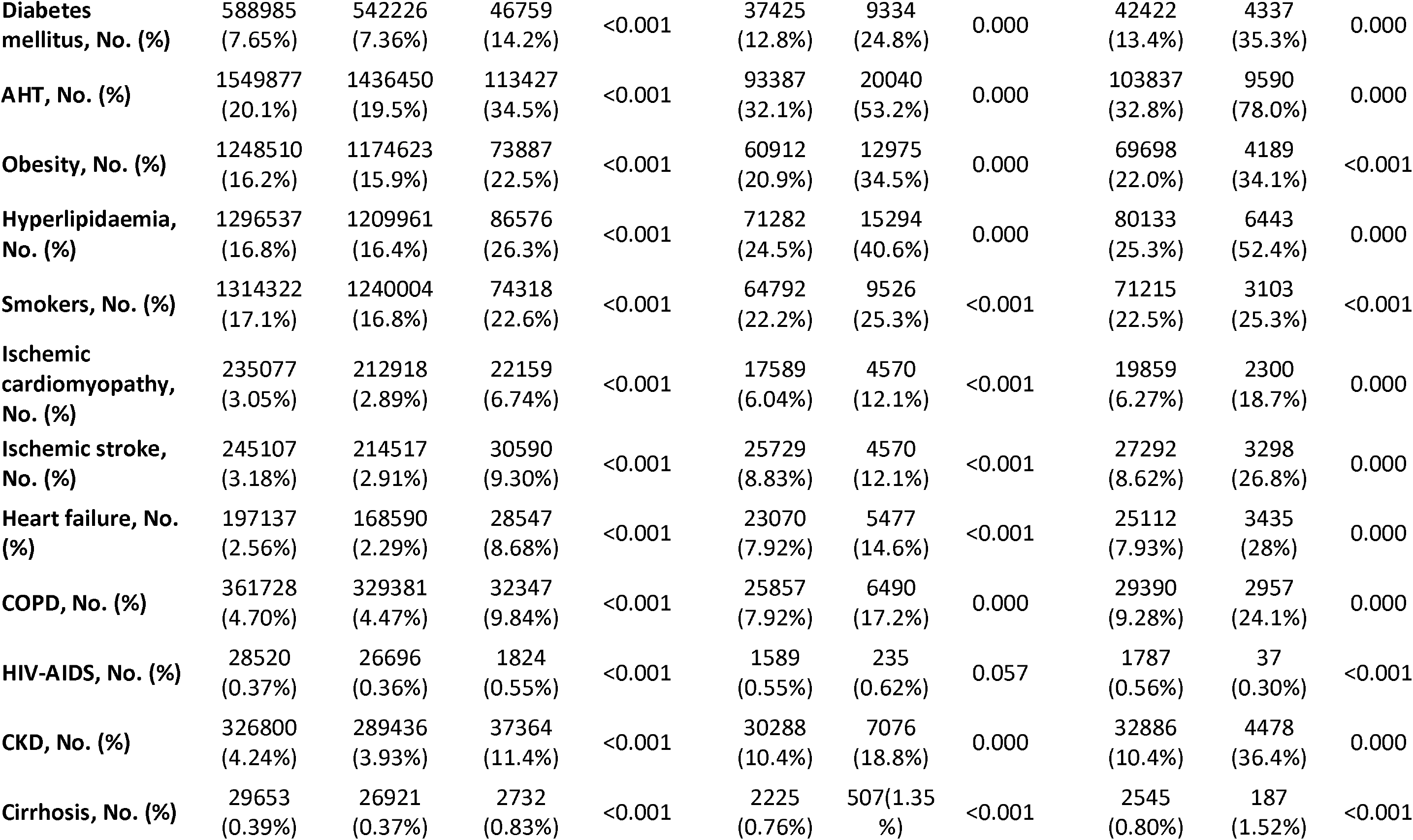
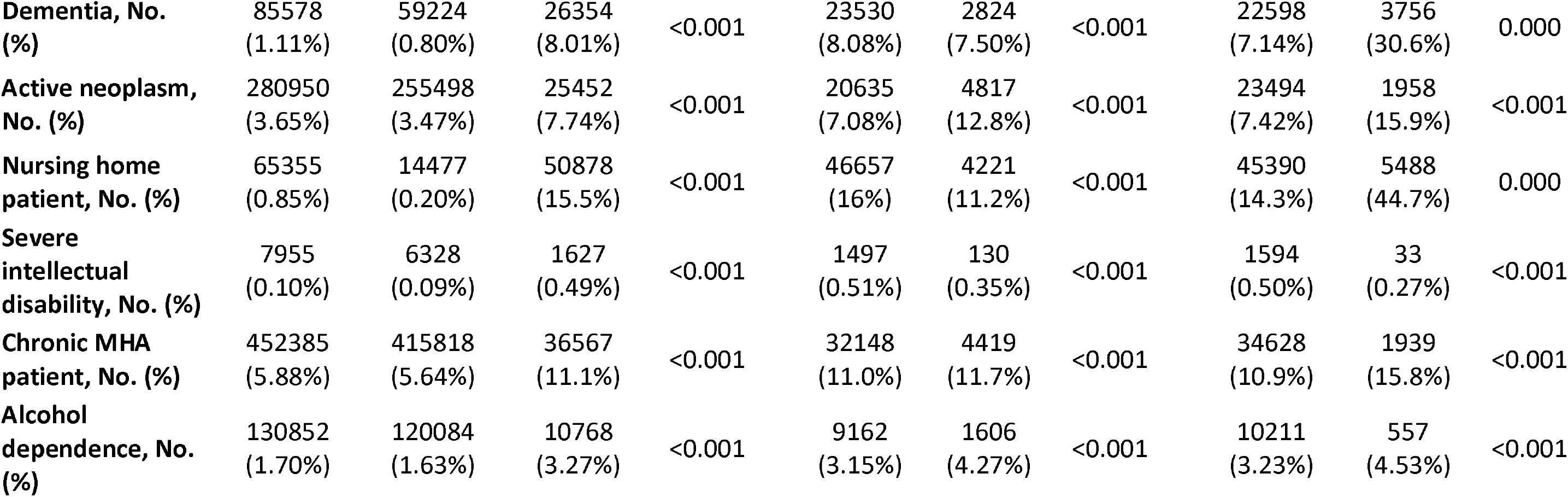
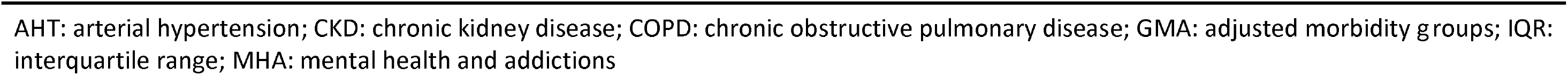
Sociodemographic variables and previous comorbidities among SARS-CoV-2 diagnosis, hospital admissions and deaths related to COVID-19 in Catalonia between 01/03/20 and 01/06/20

We used Poisson regression models to build a predictive model for infection by SARS-CoV-2 and hospitalisation and death due to COVID-19. The models were created by using a “stepwise-forward” approach based on the Akaike Information Criterion (AIC), in which a naïve model is complemented sequentially with the most relevant variables, eventually leading to the definitive model.

Finally, we used the same approach described above to perform a sensitivity analysis in PCR-positive patients only. This analysis allowed us to compare the results of both analyses and validate the predictor factors.

All analyses were performed using R statistical software, version R-4.0.0.

## Results

### Demographic and clinical characteristics

During the study period, 328 892 patients were considered to be infected with SARS-CoV-2, representing 4.27% of the entire Catalan population. Figure 1 shows the distribution of infections, hospitalisations, and deaths according to age, gender, and comorbidity burden (GMA index). The median age of this group was 53.0 years (IQR: 38.0; 74.0).

**Figure 1.**
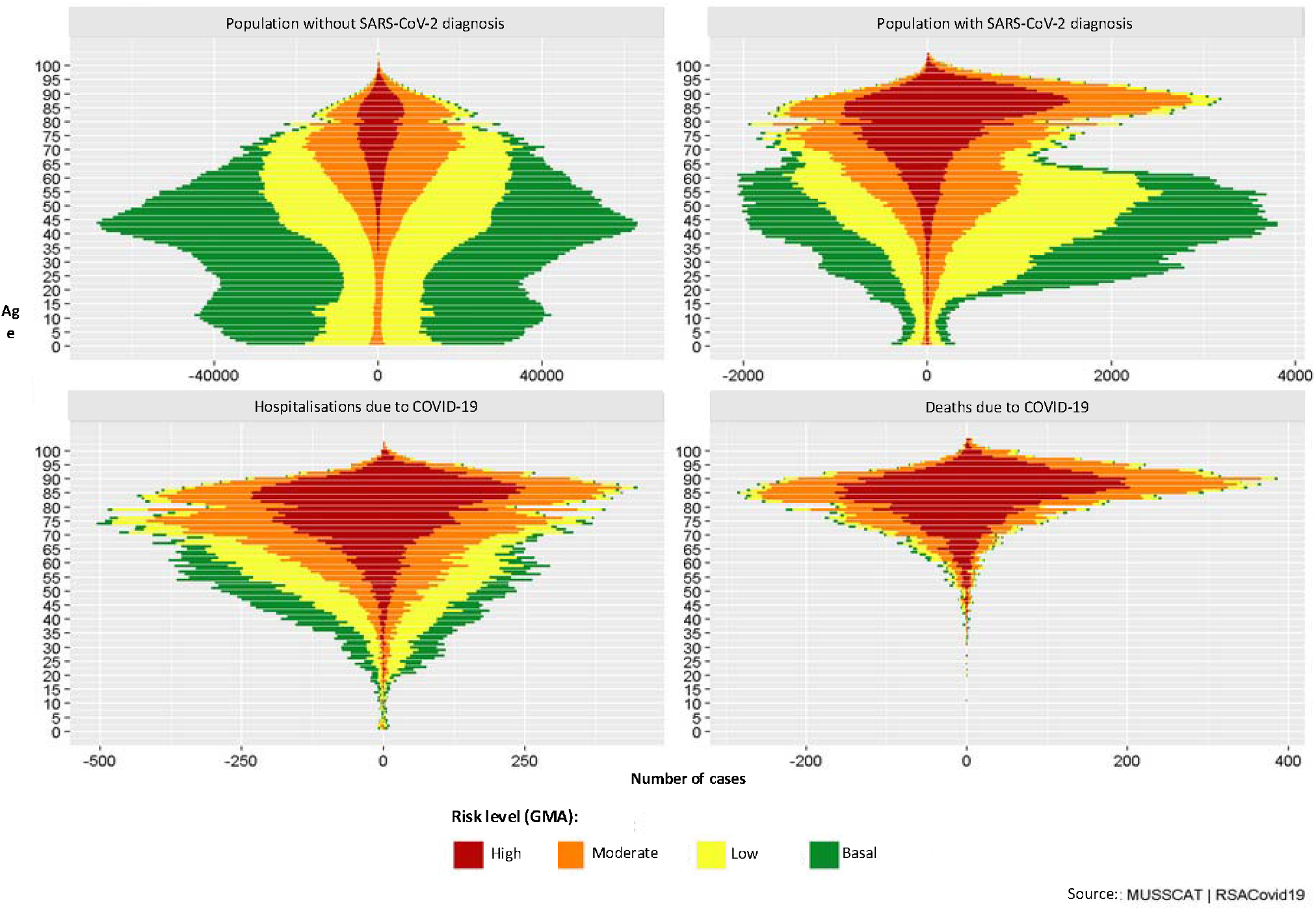
Age and sex distribution of the cases of SARS-CoV-2 diagnosis vs non-infected population and cases of hospitalisation and death by COVID-19 in Catalonia, Spain, between 01/03/2020 and 01/06/2020 GMA: adjusted morbidity groups; COVID-19: coronavirus disease 2019; SARS-CoV-2: severe acute respiratory syndrome coronavirus 2.

The group with SARS-CoV-2 infection were predominantly female (206 074, 62.7%), aged 15 to 64 years (211 383, 64.2%), of low-income status (195 641, 59.5%), and had a high burden of comorbidities according to the GMA index (54 130, 16.5%) (Table 1).

A total of 37 638 (11.4 %) patients admitted to hospitals were diagnosed as COVID-19 positive. Their median age was 68.0 years (IQR: 52.0; 81.0). These cases of hospitalisation were predominantly men (19 904, 52.9%), aged >65 years (20 882, 55.5%), and had a low individual income (23 663, 62.9%), and nearly a third had a high burden of comorbidities. The most common comorbidities were hypertension (20 040, 53.2%), hyperlipidaemia (15 294, 40.6%), and obesity (12 975, 34.5%).

During the study period, a total of 12 287 (3.73%) deaths were attributed to COVID-19, with a median age of 85 years (IQR: 77.0; 90.0). These cases of death due to COVID-19 were predominantly women (6 332, 51.2%), of low-income status (8 849, 72.0%), and almost half of them had a high burden of comorbidities according to the GMA index. Almost half of the deaths occurred in nursing home patients (5 488, 44.7%).

### Risk factors

Our predictive model showed that the main risk factors for SARS-CoV-2 diagnosis were female gender (risk ratio [RR]=1.49; 95% CI=1.48-1.50), being aged 45 to 64 years (RR=1.02; 95% CI=1.01-1.03), having a high burden of comorbidities (RR=3.03; 95% CI=2.97-3.09), having a high individual income (RR=1.06; 95% CI=1.03-1.1), being a smoker (RR=1.06; 95% CI=1.05-1.07). Residing in a nursing home was associated with a 12-fold increase in risk of infection (RR=11.82; 95% CI=11.66-11.99). Most of the comorbidities analysed individually were considered risk factors, except for hyperlipidaemia, hypertension, and obesity. Other psychiatric conditions, such as alcohol dependence (RR=1.17; 95% CI=1.14-1.20), were also associated with a higher risk of infection (Figure 2).

**Figure 2.**
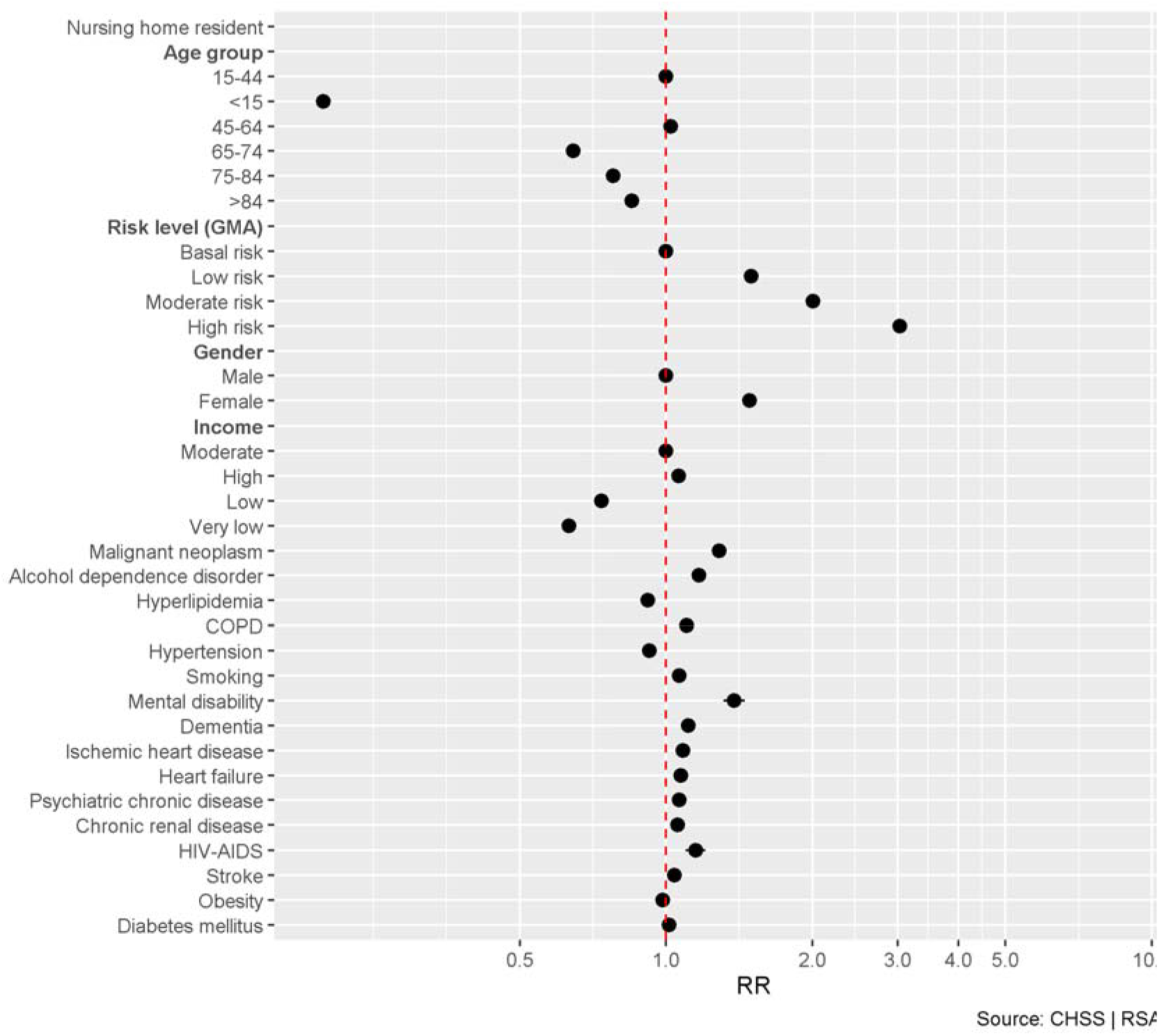
Risk ratios (shown on a log scale) and 95% confidence intervals for each potential risk factor of SARS-CoV-2 **diagnosis** from a Poisson regression model. Data collected in Catalonia, Spain, between 01/03/2020 and 01/06/2020. GMA: adjusted morbidity groups; COPD: chronic obstructive pulmonary disease; SARS-CoV-2: severe acute respiratory syndrome coronavirus 2. Gender is considering male as reference.

Our model identified the following main risk factors for hospitalisation due COVID-19: male gender (RR=1.45; 95% CI=1.43-1.48), being aged >65 years (RR=2.38; 95% CI=2.28-2.48), having a high burden of comorbidities (RR=5.15; 95% CI=4.89-5.42), having a very low individual income (RR=1.03; 95% CI=0.97-1.08), and residing in a nursing home (RR=3.97; 95% CI=3.82-4.12). The model also identified certain comorbidities, including: obesity (RR=1.23; 95% CI=1.20-1.25), COPD (RR=1.19; 95% CI=1.15-1.22), heart failure (RR=1.19; 95% CI=1.16-1.22), diabetes mellitus (RR=1.07; 95% CI=1.04-1.10), and psychiatric comorbidities (RR=1.06; 95% CI=1.03-1.10) (Figure 3).

**Figure 3.**
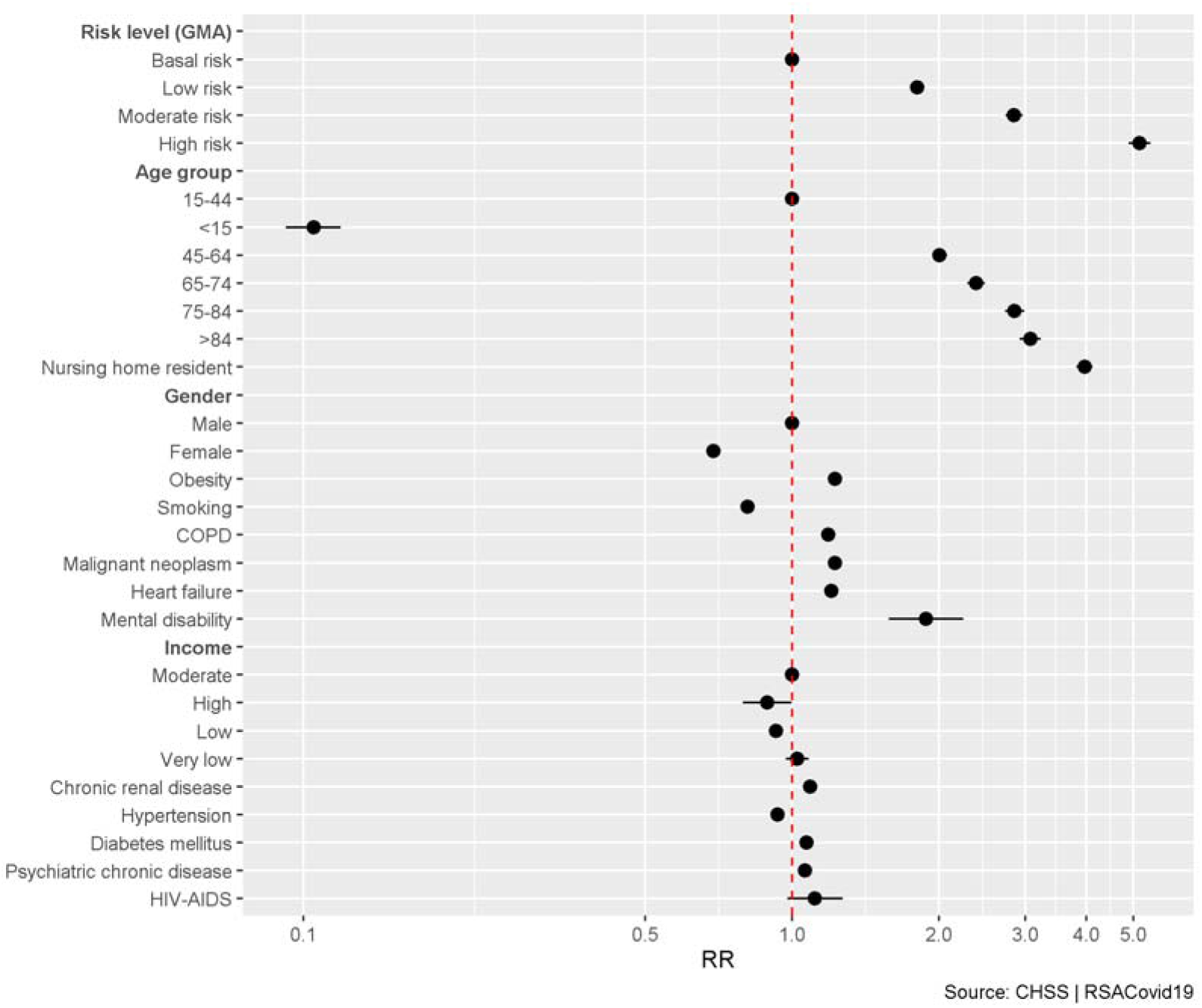
Risk ratios (shown on a log scale) and 95% confidence intervals for each potential risk factor of **hospitalisation** by COVID-19 from a Poisson regression model. Data collected in Catalonia, Spain, between 01/03/2020 and 01/06/2020. GMA: adjusted morbidity groups; COVID-19: coronavirus disease 2019; COPD: chronic obstructive pulmonary disease. Gender is considering male as reference.

Regarding deaths due to COVID-19, our model found that the most important risk factors were: male gender (RR=1.73; 95% CI= 1.67-1.81), being aged >65 years (RR=37.45; 95% CI=29.23-47.93), and residing in a nursing home (RR=9.22; 95% CI=8.81-9.65). In addition, the individual comorbidities with the highest influence on risk of death were heart failure (RR=1.21; 95% CI=1.16-1.22), CKD (RR=1.17; 95% CI=1.13-1.22), and diabetes mellitus (RR=1.10; 95% CI=1.06-1.14). Having a chronic psychiatric condition was also associated with risk of death (RR=1.15; 95% CI=1.09-1.21). Overall, the risk of death was 5-fold higher in cases with a high burden of comorbidities according to the AMG index (RR=5.25; 95% CI=4.60-6.00) (Figure 4).

**Figure 4.**
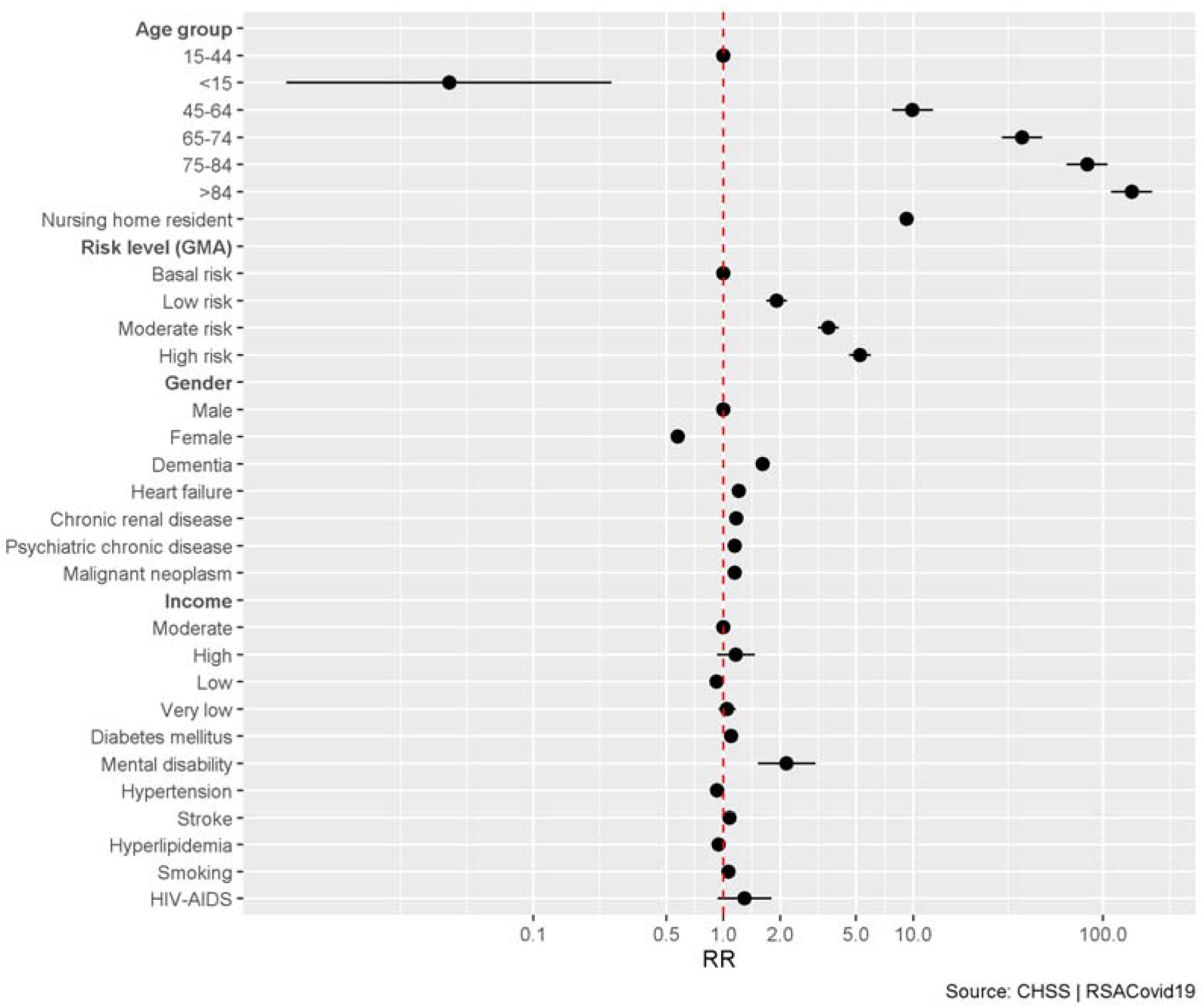
Risk ratios (shown on a log scale) and 95% confidence intervals for each potential risk factor of **death** by COVID-19 from a Poisson regression model. Data collected in Catalonia, Spain, between 01/03/2020 and 01/06/2020. GMA: adjusted morbidity groups; COVID-19: coronavirus disease 2019; COPD: chronic obstructive pulmonary disease. Gender is considering male as reference.

Our sensitivity analysis in PCR-positive patients only (53 801, 16.4%) showed that these patients had similar baseline characteristics to those in the main analysis (see supplementary table S1), although there were some differences in the risk predictors for SARS-CoV-2 infection (figure S1). Consistent with the results of the main analysis, increasing age and comorbidity burden were directly correlated with increasingly higher risk of having COVID-19, while smoking and low individual income were associated with a lower risk. The risk predictors for hospitalisation and death due to COVID-19 were similar in both analyses, with minor differences in magnitude. Very low individual income was a risk factor for death (figures S2 and S3).

## Discussion

This study used data from Catalonia, one of the regions of Spain that was most affected by COVID-19, between 1 March 2020 and 1 June 2020. By comparison with the general population, our study found that female gender and high burden of comorbidities were risk factors associated with SARS-CoV-2 infection. These subgroups of the population were therefore more likely to transmit coronavirus. Conversely, the individuals who were hospitalised or who died due to COVID-19 were mainly male, aged >65 years, or had a low-income, certain chronic conditions, or a generally high burden of comorbidities.In contrast to a previous report on the risk factors for SARS-CoV-2 infection in the UK,^6^ the risk factors in our study were female gender, being 45 to 64 years old, residing in a nursing home, and being a smoker. These differences could be due to our larger sample size, the inherent differences in the populations of different countries, and the differences in diagnostic criteria for SARS-CoV-2 infections. However, our sensitivity analysis in PCR-positive patients only, age was positively associated with risk, while smoking and low individual income were associated with lower risk. This last finding was also reported elsewhere^6^, and was suggested to be due to confounding factors, such as the effect of active smoking on the sensitivity of PCR-tests. Moreover, healthcare workers are considered a high-risk population because of their exposure,^24^ and nursing homes in Catalonia were a very active focus of SARS-CoV-2 infection, especially since more than half of residents and staff were asymptomatic carriers.^8^ Thus, in our study, the fact that female gender was a risk factor for infection could be due to the overrepresentation of women in care activities.

Based on the results of previous models, “stratify-and-shield” strategies have been proposed to enhance protection among vulnerable population groups, while easing restrictions on the rest.^28–33^ These approaches aim to identify groups at high risk of dying from COVID-19 and then shield them against infection; this strategy is pragmatic because unshielded individuals with a low risk of morbidity and mortality will not overload hospitals’ ICUs.^34^ However, this approach only considers risk factors for hospitalisation or death by COVID-19, whereas we believe it is also important to identify individuals with the highest risk of SARS-CoV-2 infection. This would also allow us to devise and apply prevention and care strategies to protect them.

In our study, the risk factors for hospitalisation and death due to COVID-19 included male gender and being aged 65 years and over. Hospitalisation was also found to be associated with having a very low individual income, or certain comorbidities, such as obesity, COPD, and heart failure. Death due to COVID-19 was associated with residing in a nursing home, heart failure, CKD, and diabetes mellitus. Most of these results are consistent with those of previous reports.^1,9–15^

Our study also considered psychiatric comorbidities, and found these also to be a risk factor for hospitalisation and death. This is important because neuropsychiatric disorders (including mental health, addictions or cognitive impairment) are usually not taken into account in studies, despite its relationship with comorbidity, dependence and polypharmacy, among other aspects due to COVID-19 that may be associated to poor prognosis. Similarly, socioeconomic status has not been consistently assessed in relation to this disease, but our results indicate that it is an important parameter that should be examined systematically. Socioeconomic conditions may influence differently if we analyze the risk of acquiring the infection rather than to analyze the risk of hospitalization or death, where the low level income is associated with a worse prognosis. Furthermore, socioeconomic status, may be a changing risk transmission factor depending on the phase of the epidemic: it may be different in the virus introduction phases (high level income is associated with tourist or business trips or attendance to mass recreational or cultural events, in phases in which there are no restrictions), than when there is already community spread (when living or working conditions associated with low level income may facilitate virus transmission).

Our model also determined that CKD was a relevant comorbidity in patients who died of COVID-19; this comorbidity has not been considered in previous extensive studies on coronavirus risk factors for death. Besides analysing individual comorbidity factors, we also used the GMA comorbidity index. GMA is a validated index for grouping morbidity that is comparable to others in the field and has been used to stratify the population and identify target populations in previous studies.^35^ We found that those individuals classified as having a high burden of comorbidities were either at higher risk for being considered infected, hospitalised, or dying from COVID-19. Therefore, in our study, the burden of comorbidities calculated by the GMA index was a risk factor that offered a better explanation of these risks than the individual comorbidities.

In addition, our sensitivity analysis identified similar risk factors in PCR-positive patients, thus validating the results of our main analysis.

### Implications and recommendations

The risk factors predicted by our model support alternative actions with public health and clinical practice implications. Mitigation strategies should be based on when (contention vs mitigation), where, and who needs to be protected. The latter is crucial, as protecting, for instance, front-line healthcare workers, both in hospitals and in nursing homes, would help avoid the spread of SARS-CoV-2, and would thus shield the population groups with the highest risk of hospitalisation, complications, and death related to COVID-19. Therefore, it is imperative that strategies to control the pandemic don’t focus only in identify the groups most at risk due to their comorbidity, but also to take into account the socioeconomic circumstances and gender (the jobs where there is closer contact with the most vulnerable population to have a severe COVID, are mainly carried out by the female gender), prioritizing in these groups public health measures to mitigate SARS-CoV-2 transmission. In addition, according to our results, mental health, comorbidity burden, and individual income are factors that should also be taken into account in further studies related to COVID-19 risk factors.

### Strengths and limitations

In this study, data were collected from the Catalan Health Surveillance System, a large healthcare database that includes a myriad of information on Catalan residents. This broad array of data, including an index for measuring the comorbidity burden, allowed us to gain a global perspective of the issues discussed, as opposed to working with just a very specific group (e.g. hospitalised patients with COVID-19). This database includes sociodemographic and other comorbidities that have not been prioritized in previous studies, and that have clearly been shown to play a role in SARS-CoV-2 transmission, and thus in disease control.

Nevertheless, a limitation of this study is its descriptive nature, although we were indeed able to build a predictive model. A major drawback was the lack of PCR confirmation for most SARS-CoV-2 infected patients: the scarcity of tests during the study period precluded an extensive robust validation of all results. In addition, the fast-changing epidemiologic definitions of COVID-19 also worked against our analysis. Finally, since ours was a near-real-time analysis, hospitalisation notifications are likely to have been underestimated.

## Conclusions

This study of a large cohort including the entire population of Catalonia, a region of Spain, has provided predictive models to identify risk factors for SARS-CoV-2 infection, hospitalisation, and death. According to our model, the main risk factors for infection were female gender, being 45 to 64 years old, residing in a nursing home, and having a high burden of comorbidities. The latter might be a better risk factor than comorbidities considered individually. It is important to precisely define these risks, in order to design interventions aimed at transmission control, rather than focusing only on the population at risk of death. In addition, we found that most risk factors for hospitalisation and death due to COVID-19 were similar to those previously reported. However, specific risk factors that had not been described to date included CKD, comorbidity burden, mental health, and individual very low income In order to avoid overwhelming the health system, it is essential to identify groups with low risk of hospitalisation or death, but with a high risk of transmitting the disease to the vulnerable population. Furthermore, it would be recommended to repeat this evaluation during the development of the epidemic, because the risk factors for transmission (like socioeconomic income) may be different depending on the spread phases. Future COVID-19 studies should examine the role of gender, the burden of comorbidities, and socioeconomic status in disease transmission, and should determine its relationship to workplaces, especially healthcare centres and nursing homes.

## Data Availability

the data will be made available to investigacional purpose with investigator group support, after approval of a proposal, with a signed data access agreement.

http://aquas.gencat.cat/.content/Enllac/factors-risc-mortalitat-covid19-hospitalitzats.html

http://aquas.gencat.cat/.content/Enllac/factors-pronostics-hospitalitzacio-covid19.html

## Acknowledgements

The authors thank Matias Rey-Carrizo PhD and Gavin Lucas PhD at www.thepapermill.eu for providing editorial support.

## SUMMARY BOXES

### What is already known on this topic

Several studies have reported the risk factors for hospitalisation and death due to coronavirus disease 2019 (COVID-19).

Some of these risk factors have been found to differ between countries.

There is limited evidence on the specific risk factors for infection by severe acute respiratory syndrome coronavirus 2 (SARS-CoV-2).

### What this study adds

In our setting (Catalonia, a region of Spain with 7 699 568 inhabitants), the risk factors for SARS-CoV-2 infection are female gender, high burden of comorbidities, and residing in a nursing home.

The risk factors for hospitalisation and death due to COVID-19 are male gender, being older than 65 years, and having a high burden of comorbidities, including neuropsychiatric disorders, and very low individual income.

Interventions to mitigate SARS-CoV-2 infection should consider the population with the highest risk of transmitting the disease, not only those at risk of death.

